# The Clinical Utility of the BMD-related comprehensive Genome-wide polygenic score in identifying individuals with a high risk of osteoporotic fractures

**DOI:** 10.1101/2022.11.16.22282416

**Authors:** Xiangxue Xiao, Qing Wu

## Abstract

**Purpose:** This study sought to construct genome-wide polygenic scores for femoral neck and total body BMD and to estimate their potential in identifying individuals with a high risk of osteoporotic fractures.

**Methods:** Genome-wide polygenic scores were developed and validated for femoral neck and total body BMD. We externally tested the PGSs, both by themselves and in combination with available clinical risk factors, in 455,663 European ancestry individuals from the UK Biobank. The predictive accuracy of the developed genome-wide PGS was also compared with previously published restricted PGS employed in fracture risk assessment.

**Results:** For each unit decrease in PGSs, the genome-wide PGSs were associated with up to a 1.17-fold increased fracture risk. Out of four studied PGSs, ***PGS_TBBMD***_**81**_ (HR: 1.03; 95%CI 1.01-1.05, p=0.001) had the weakest and the ***PGS_TBBMD***_***ldpred***_ (HR: 1.17; 95%CI 1.15-1.19, p<0.0001) had the strongest association with an incident fracture. In the reclassification analysis, compared to the FRAX base model, the models with, ***PGS_FNBMD***_**63**_, ***PGS_TBBMD***_**81**_, ***PGS_FNBMD***_***ldpred***_, and ***PGS_TBBMD***_***ldpred***_ improved the reclassification of fracture by 2% (95% CI, 1.5% to 2.4%), 0.2% (95% CI, 0.1% to 0.3%), 1.4% (95% CI, 1.3% to 1.5%), and 2.2% (95% CI, 2.1% to 2.4%), respectively.

**Conclusions:** Our findings suggested that an efficient PGS estimate enables the identification of strata with up to 1.5-fold difference in fracture incidence. Incorporating PGS information into clinical diagnosis is anticipated to increase the benefits of screening programs in the population level.

## Introduction

Osteoporosis is an age-related, devastating bone disease characterized by low bone mineral density (BMD) and structural deterioration of bone tissue [1], resulting in an increased risk of fracture. As the world population ages rapidly, bone fracture is becoming a major public health issue. Each year, osteoporosis is responsible for more than 8.9 million fractures globally, of which more than 1.5 million occur in the United States [2]. In 2025, osteoporotic fractures are projected to increase to over 3 million in the US [3]. The increasing fracture incidence renders early identification and preventive intervention a vital goal.

Several fracture predictive tools have been developed in recent years. In the United States, the Fracture Risk Assessment Tool (FRAX) is the most widely used fracture prediction tool, which is well-established and validated to predict 10-year probabilities of major osteoporotic fracture (MOF) and hip fracture (HF) on the basis of 12 clinical risk factors [4]. However, the performance of FRAX in discriminating fracture and non-fracture cases is too often unsatisfactory, which certainly indicates that there is still room for improvement [5-7].

The predisposition to osteoporotic fracture is attributable to the complex interaction between genetic and non-genetic factors [8]. As a major determinant of fracture risk, BMD measured by dual-energy X-ray absorptiometry (DXA) has been proven to be highly heritable [9-13] and has thus been widely investigated in Genome-wide association studies (GWAS) [14-16]. Numerous BMD-associated genetic variants, mainly single nucleotide polymorphism (SNPs), have been discovered in the past decade [15-17]. As a result, the polygenic score (PGS), calculated according to GWAS summary statistics and an individual’s genotype profile, is often used to quantify the genetic propensity of individuals to a disease/trait [18].

Prior studies have demonstrated the potential use of BMD-decreasing PGS in predicting fracture risk; however, they provided only limited predictive power [19-22]. A PGS based on 62 femur neck-related SNPs revealed a hazard ratio (HR) of 1.20 for incident fracture per one standard deviation (SD) increase [23]. Another study derived a similar PGS from 63 BMD-related SNPs was also reported having significant association with fracture risk in adults [24]. However, previously published genetic risk scores included genetic variants restricted to those that reached genome-wide significant levels (p < 5*10^−8^). Due to the polygenic nature of BMD, previously established “restricted PGSs” were not able to sufficiently capture the underlying genetic predisposition, thus failing to provide a comprehensive assessment of genomic information in fracture risk prediction. PGSs calculated from millions of variants across the genome and accounting for linkage disequilibrium (LD) between variants was proven to outperform traditional PGS in the risk prediction of several diseases, such as cardiovascular disease, type II diabetes, and breast cancer [25-27]. However, whether a novel BMD-related genome-wide PGS derived from an improved PGS algorithm would significantly increase the predictive power of the genetic components in fracture prediction remains unclear. Therefore, we aimed to build more robust and generalizable genome-wide PGSs for BMD to provide a more comprehensive fracture risk evaluation. We compared the accuracy of the genome-wide PGS with previously published PGS in fracture risk assessment. We also aimed to assess the added value of PGS beyond FRAX in fracture prediction. We hypothesized that genome-wide PGSs would outperform previously published “restricted PGS” in assessing fracture risk and that combining genome-wide PGS with FRAX could better identify individuals at high risk of osteoporotic fracture.

## Method

### Study cohort

UK Biobank (UKB) is a large-scale, population-based observational study consisting of 502,617 individuals aged 40-69, recruited from across the United Kingdom between 2006 and 2010 [28]. A total of 488,251 participants were genotyped using Affymetrix arrays [29]. The genotype data were quality controlled and imputed using the Haplotype Reference Consortium [30]. At recruitment, a standardized socio-demographic questionnaire, medical history, and other lifestyle factors were collected. Individual records were linked to the Hospital Episode Statistics (HES) records and the National Death and Cancer Registries. Compared to the general population, the UKB participants were healthier, less obese, and less likely to smoke and drink alcohol [31]. Since the PGSs were derived based on predominately White GWAS participants and the people of non-European ancestry comprised only a small proportion of the UKB, we restricted the analysis to 452,936 white British individuals so as to analyze individuals with a relatively homogeneous ancestry.

### Fracture event ascertainment

Fracture cases were identified through the Hospital Episodes Statistics linked through NHS Digital, with a hospital-based fracture diagnosis irrespective of mechanism within the primary (data field #41202; n= 435,968) or secondary (n= 435,972) diagnosis field. Fractures of the skull, face, hands, and feet, as well as pathological fractures due to malignancy, atypical femoral fractures, periprosthetic, and healed fracture were excluded from the analysis. The incident fracture cases were defined as having the date of ICD-10–identified fractures after the initial assessment visit.

### Ascertaining conventional risk factors

Age, sex, height, weight, body mass index (BMI), previous fracture, current smoking status, glucocorticoid use, rheumatoid arthritis, and secondary cause of osteoporosis (Type 1 diabetes and menopause before age 45 years) were ascertained from the initial assessment visit. Previous fractures were defined as those reported by questionnaire at enrollment or from ICD-10 codes that occurred before the baseline visit. Gender was self-reported and verified by genotype, and Individuals with discordant sex between self-report and genotype were excluded.

### Data processing and quality control

Genotyping of the UKB samples was performed using Affymetrix, UK BiLEVE Axiom, and the Affymetrix UKB Axiom array. The Wellcome Trust Centre for Human Genetics performed the genotype imputation using the Haplotype Reference Consortium (HRC) and the UK10K haplotype resources, which yielded a total of 96 million imputed variants. Quality control was performed for the UKB genotype data: SNPs with minor allele frequency less than 0.1%, were missing in a high fraction of subjects (>0.01), and have Hardy-Weinberg equilibrium p-value < 1*10^−6^ were removed. Individuals who have a high rate of genotype missingness (> 0.01) were also excluded from PGS construction. After quality control, a total of 11.5 million variants were retained for analysis.

### Polygenic score tuning

The summary statistics of two comprehensive GWA studies conducted among European predominantly cohorts for femoral neck BMD [16] and total body BMD [14] were used to derive PGSs. UKB samples were not included in any of the two discovery GWASs. The UKB dataset was split into a tuning set (n=3,000) and a testing set (n=452,936). For the tuning set, we randomly selected 1000 prevalent fracture cases and 2000 non-fracture cases of European ancestry. A set of candidates PGSs was derived for each trait by using the Pruning and Thresholding (P+T) method and the LDPred2 computational algorithm in the tuning set.

The P+T method PGSs were built using a *p*-value and linkage disequilibrium-driven clumping procedure in PLINK 1.90b. Twenty-four candidate PGSs were identified as having combinations of the *p*-value (1.0, 0.5, 0.05, 5 × 10^−4^, 5 × 10^−6^, and 5 × 10^−8^) and r^2^ (0.2, 0.4, 0.6, and 0.8) thresholds for each trait.

The LDPred2 computational algorithm was used to generate seven candidate PGSs for each trait. Based on seven hyper-parameter values of ρ (1, 0.3, 0.1, 0.03, 0.01, 0.003, and 0.001), seven sets of candidates PGSs were generated using the LDPred2 computational algorithm grid mode. Each set of PGSs tested a grid of hyper-parameter values, where 102 combinations of hyper-parameters ρ (the proportion of causal variants) and *h*^2^ (the SNP heritability) were tuned. For each ρ value, we chose the best model according to the Z-score from the regression of the fracture by the PGS, with age, sex, and BiLEVE/UKB genotyping array and the first four principal components (PCs) being adjusted for. The PGS construction was restricted to the HapMap3 variants only as LDpred2 suggested [33].

Together there were 31 candidate PGSs that have been derived. The association between PGS and fracture was further evaluated in odds ratios (OR) per standard deviation of PGS using logistic regression adjusted for age, sex, and BiLEVE/UKB genotyping array and the first four principal components (PCs). The femoral neck BMD-related PGS (*PGS_FNBMD*_*ldpred*_) and total body-related PGS (*PGS_TBBMD*_*ldpred*_) with the maximum predictive ability (AUC) with fracture were determined to be the best-performing ones and were carried forward into subsequent analyses in the independent UKB testing set. For femoral-neck BMD and total body BMD, p thresholds of 0.03 and 0.13, respectively, provided the most optimal discrimination of fracture cases and controls and were chosen to derive the genome-wide PGSs in the UKB testing set for the subsequent analyses. We additionally calculated two previously published femoral neck and total body-related PGSs from Estrada et al. (*PGS_FNBMD*_63_) and Xiao et al., (*PGS_TBBMD*_81_) in the UKB testing set so as to compare the predictive value of the genome-wide PGS with the “restricted PGS” in assessing fracture risk (**Figure 1**). Since the PGSs were BMD-related, greater PGS is associated with higher BMD and lower fracture risk.

**Figure 1.**
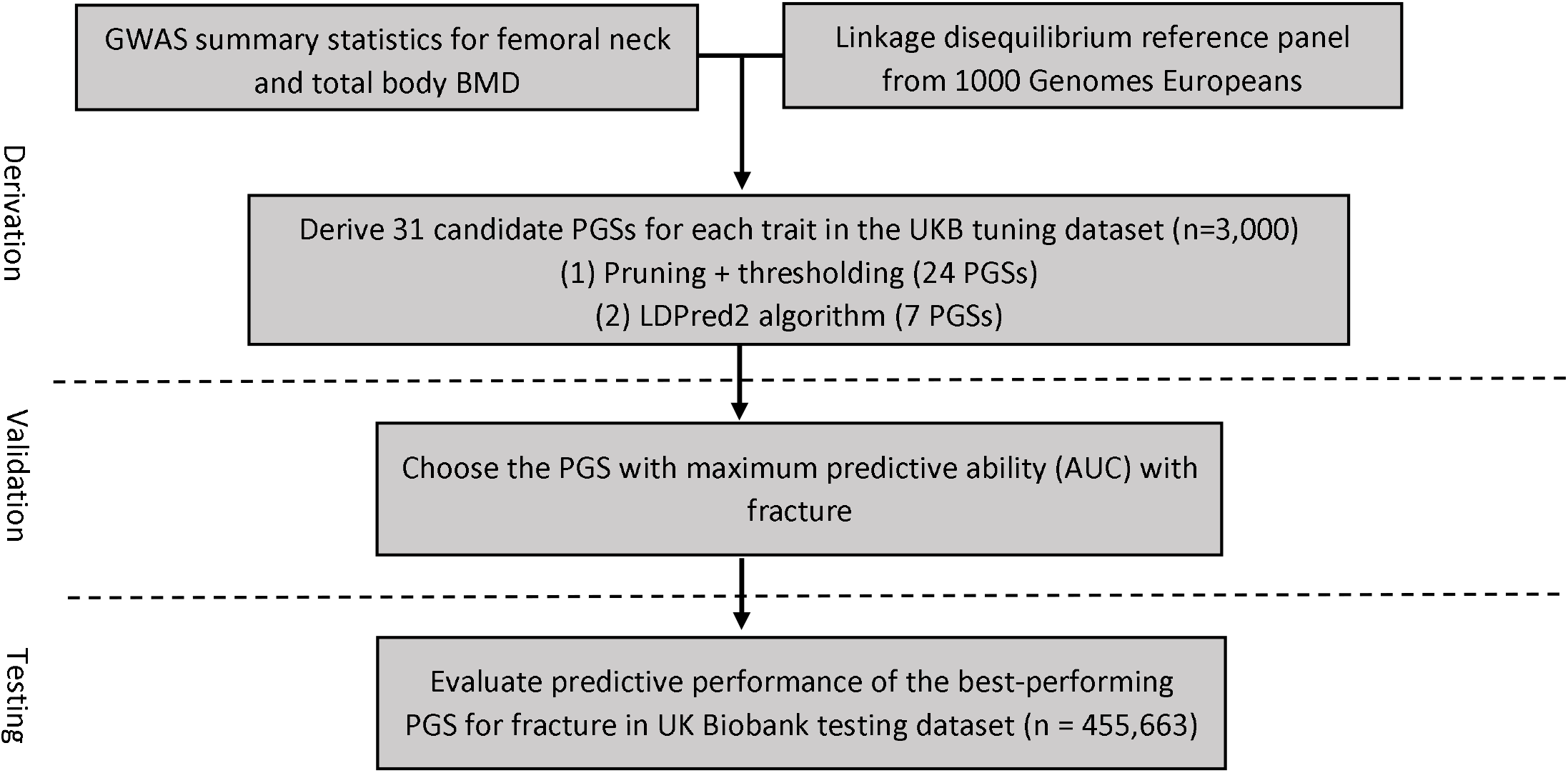
Study Design and Workflow.

### Statistical Analysis

Demographic and baseline clinical characteristics of the UKB testing set are presented as mean ± standard deviation (SD) for continuous variables and frequencies (%) for categorical variables. All PGSs were standardized to zero-mean and unit-variance. The primary outcome was incident fracture that occurred after the baseline visit.

To gauge the potential clinical impact of PGSs, we binned the UKB testing set into 100 groupings based on the percentile of the PGSs and determined the prevalence of fracture within each bin, stratified by sex. The predicted probability of incident fracture based solely on PGSs was also examined by gender. We additionally compared the observed risk gradient with the PGS predicted risk across percentile bins. For each individual, the 10-year predicted probability of disease was calculated using simple logistic regression model includes PGS only. The predicted prevalence of disease within each percentile bin was calculated as the average probability of all individuals within that bin predicted solely by PGS. To illustrate the different cumulative incidence of fracture in individuals with distinct genetic predispositions, we grouped individuals according to different quantile ranges of PGSs: ≤1%, 1-5%, 5-20%, 20-40%, 40-60%, 60-80%, 80-95%, 95-99%, and >99%. The cumulative incidence of fracture by each PGS group was then derived using the cumulative incidence function (CIF), with the competing mortality risk accounted for.

The association between incident fracture risk and each PGS was first assessed using multiple logistic regression models. The discriminatory accuracy of each model was also evaluated using the c-index. Next, we used the Cox proportional hazard modeling to estimate HRs of PGSs on incident fractures. The Cox proportional hazard model’s proportionality assumption was visually inspected beforehand using the Schoenfeld Residual test [34], and the linearity assumption was checked using the Martingale Residual test [35]. The UKB testing set satisfied both the proportional hazards and linearity assumptions. Additionally, we examined fracture incidence according to the PGS category in the UKB testing set. We compared the effect of top percentiles (1%, 5%, 10%, and 20%) with the remaining percentiles (99%, 95%, 90%, and 80%) of each PGS. using Cox proportional hazard models. The predictive performance of each PGS was also assessed using the C-index. All analyses were adjusted for age, sex, and the first four principal components (PCs).

We also investigated the predictive value of PGS beyond the existing fracture assessment tool. The association between PGS with fracture risk, adjusted for the FRAX risk factors, including age, body weight, height, previous fracture, current smoking, glucocorticoids, and rheumatoid arthritis, was assessed using Cox proportional hazard models. The model with only FRAX risk factors included was set as the base model. In total, five models were formulated as follows: Model 1 – FRAX base model; Model 2 – FRAX + *PGS_FNBMD*_63_; Model 3 – FRAX + *PGS_TBBMD*_81_ ; Model 4 – FRAX + *PGS_FNBMD*_*ldpred*_ ; and Model 5 – FRAX + *PGS_TBBMD*_*ldpred*_. The magnitude of the association between each PGS and fracture risk was assessed by the HRs and its corresponding 95% confidence intervals. In addition, net reclassification improvement (NRI) comparing the nested models was calculated separately for individuals with and without fractures. We designated “high risk” as predicted MOF risk ≥ 20% and “low risk” as predicted MOF risk < 20%, based on the National Osteoporosis Foundation recommended fixed intervention cutoff. The Integrated discrimination improvement (IDI) was also calculated to incorporate both the direction of change in the calculated risk and the extent of change. All statistical analyses were conducted using R version 4.0.3 software and SAS.

## Results

### Characteristics of the UKB testing set

The characteristics of the UKB participants in the testing set (N=455,663) are shown in **Supplementary Table 1**, comprising 17,351 fracture cases and 441,196 non-fracture cases in total. There were 5,720 prevalent fracture cases at the time of recruitment and 11,649 incident cases of fracture during a mean follow-up of 6.2 years. In the UKB testing set, the four PGSs were moderately correlated, with correlation coefficients ranging from -0.03 to 0.43.

### Fracture risk by PGS groups

In the UKB testing cohort, a lower PGS, which predicts a lower BMD, was associated with higher fracture risk. Our results showed that, for both men and women, the *PGS_FNBMD*_63_, *PGS_FNBMD*_*ldpred*_, and *PGS_TBBMD*_*ldpred*_ percentile among fracture cases were higher than among healthy controls. The distribution of *PGS_TBBMD*_81_; however, it did not show a big difference between fracture cases and non-cases (**Figure 2A & Supplementary Figure 1A**). Similarly, the predicted probability of incident fracture was significantly higher among women than among men, and a sharp decrease can be observed in the right tail of the *PGS_FNBMD*_63_, *PGS_FNBMD*_*ldpred*_, and *PGS_TBBMD*_*ldpred*_ distributions. Individuals with higher BMD-related PGS have a lower risk of fracture **(Figure 2B & Supplementary Figure 1B)**. Based only on the PGSs, the shape of the observed risk gradient was consistent with predicted risk, except for PGS_tbbmd81 **(Figure 2C & Supplementary Figure 1C.)**. The crude 10-year cumulative fracture incidence by nine PGS groups was shown in Figure 5. With competing mortality risk accounted for, significant differences were observed across *PGS_FNBMD*_63_, *PGS_FNBMD*_*ldpred*_ and *PGS_TBBMD*_*ldpred*_ groups (p<0.0001). The crude fracture incidence was significantly higher among individuals with low PGS **(Figure 3)**.

**Figure 2.**
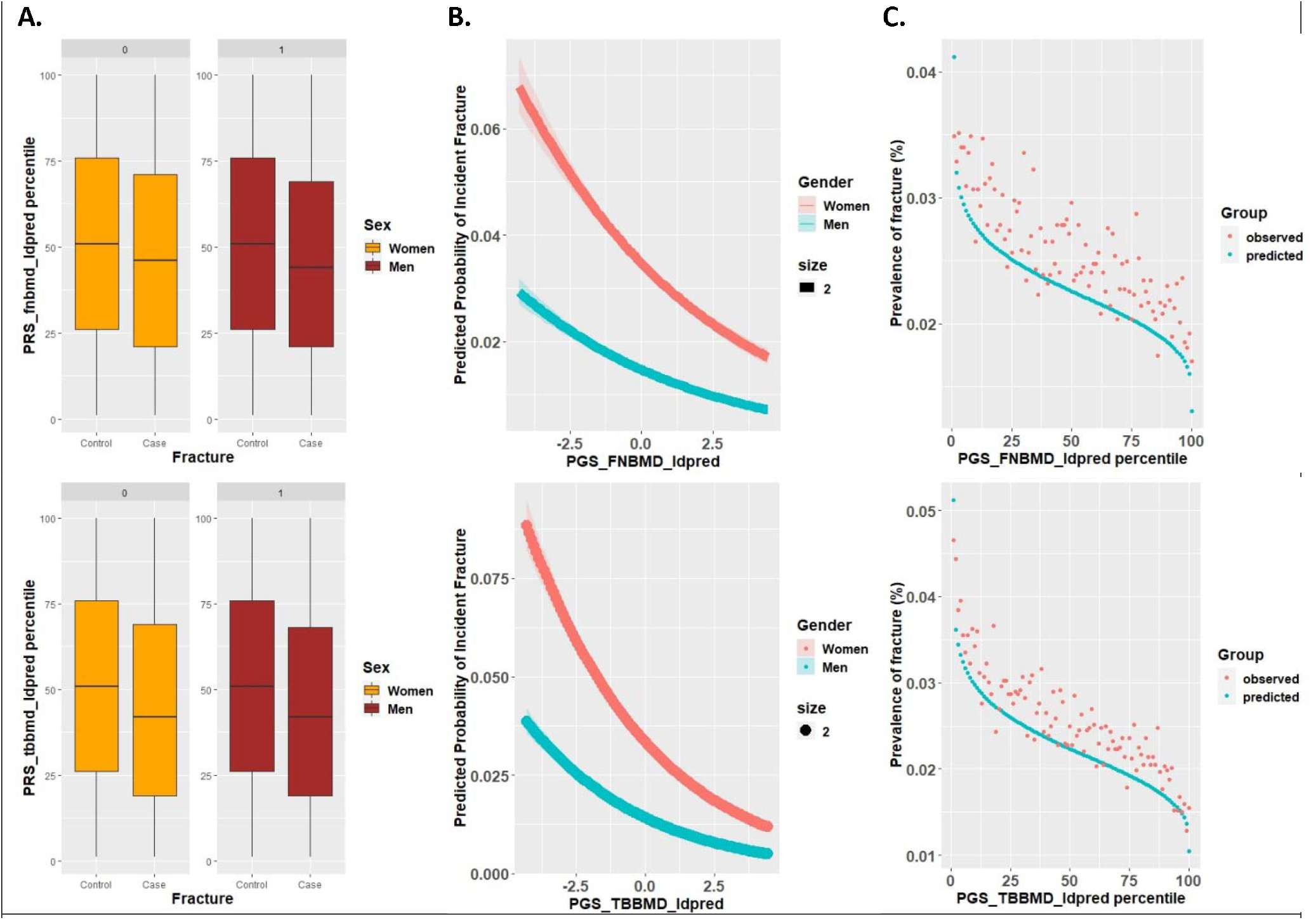
Risk for Incident Fracture According to Genome-wide PGSs. **(A)** PGSs percentile among fracture cases versus controls in the UK Biobank testing set. Within each boxplot, the horizontal lines reflect the median, the top and bottom of each box reflect the interquartile range, and the whiskers reflect the maximum and minimum values within each group. **(B)** Predicted Probability of Incident Fracture by PGSs: Risk gradient for fractures according to the PGS percentiles. 100 groups of the testing dataset were derived according to the percentile of each of the four PGSs. **(C)** Predicted versus Observed prevalence of incident fracture according to PGS percentiles.

**Figure 3.**
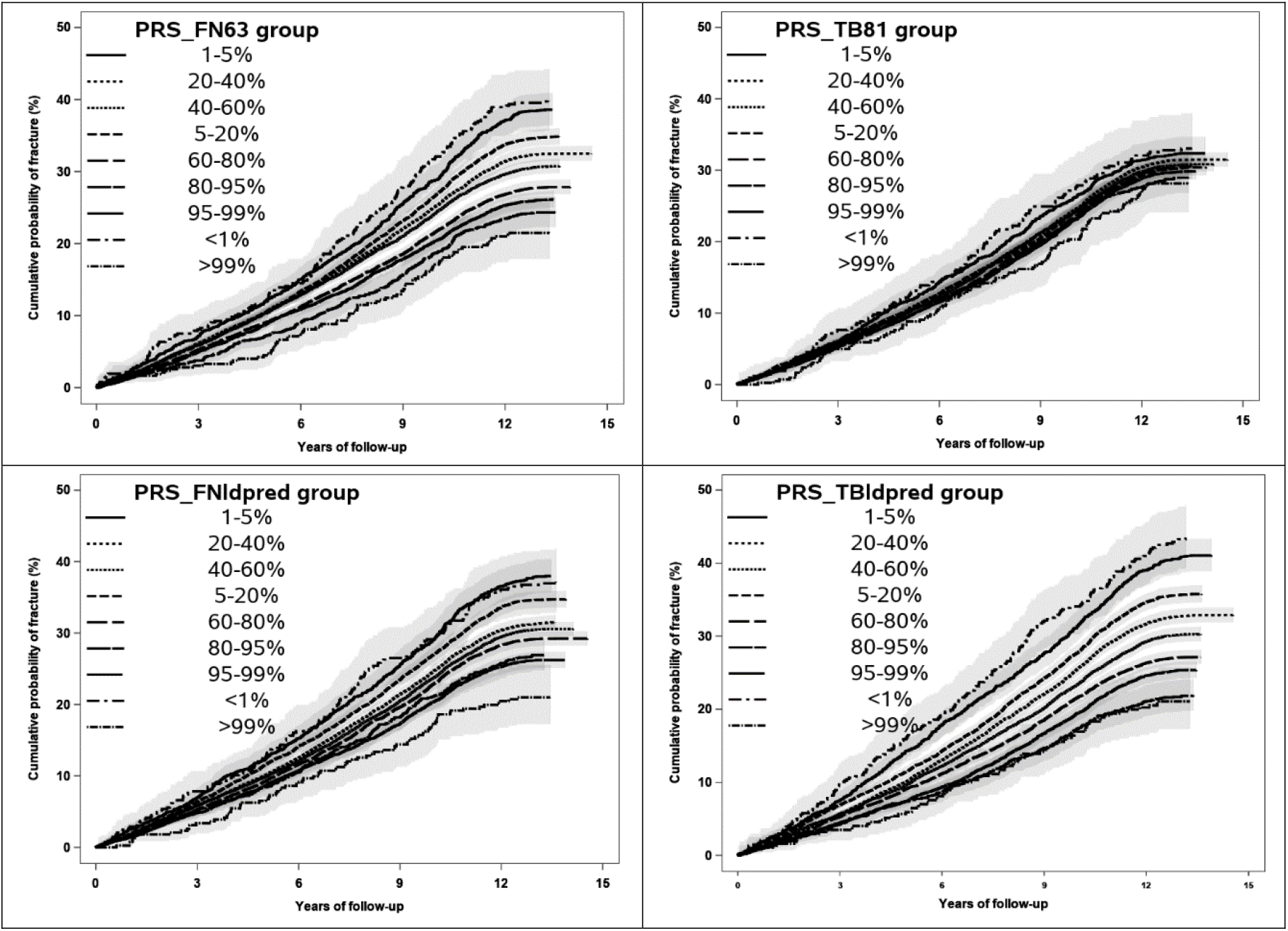
Cumulative Incident Function Plot for Fracture According to Decile of the Genome-Wide Polygenic Score (PGS) in UKB Testing Set. Shaded Regions Denote 95% Confidence Intervals.

### PGSs association with incident fractures

Multiple logistic regression results show that, in the UKB testing set, each of the four GPSs was strongly associated with incident fracture (p<0.0001), with an OR ranging from 1.03 to 1.27. A comparison of the genome-wide PGS with previously published PGS from Estrada et al., () and Xiao et al. () in the UKB testing set is given in **Figure 4A**, showing that the genome-wide PGS of total body BMD had a substantially greater association with fracture risk in terms of OR, whereas the genome-wide PGS of femoral neck BMD () didn’t show a significantly higher association with fracture compared to the restricted PGS (). For total body BMD related PGS, the genome-wide PGS () outperformed the restricted PGS () with OR estimates per standard deviation decrease at 1.03 (95% CI, 1.01 – 1.05) and 1.27 (95% CI, 1.25 – 1.30) of the and, respectively.

**Figure 4.**
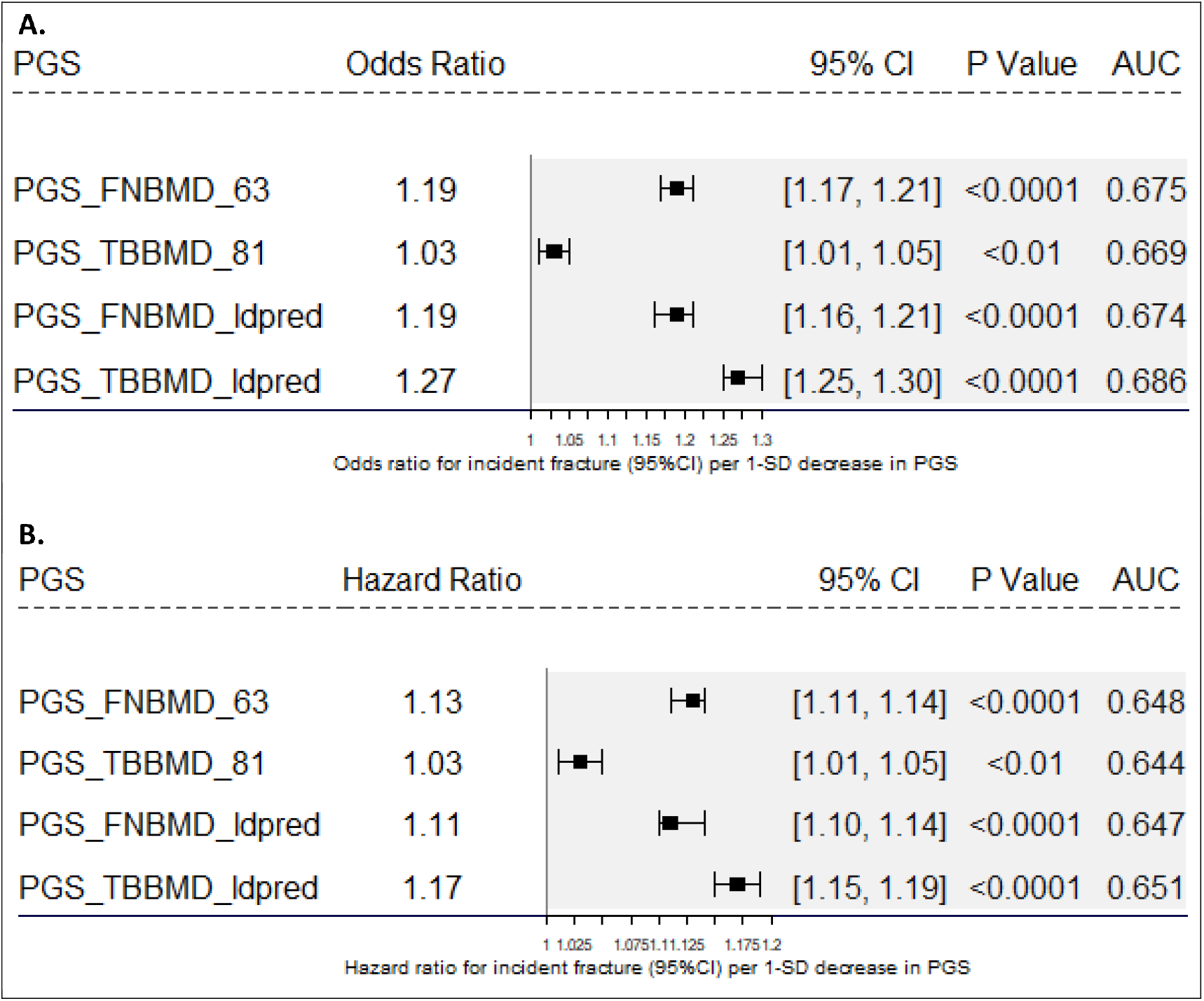
Relative Performance of Individual Polygenic Scores (PGS) for Fracture. 4A: Results from Cox Proportional Hazard Models; 4B: Results from Multivariate Logistic Regression Models.

The Cox proportional hazard regression results showed attenuated but significant associations between each PGS and the fracture risk. For every one unit decrease of PGSs, the restricted PGS and the genome-wide PGSs were associated with up to 1.13-fold and 1.17-fold increased fracture risk, respectively. Out of four studied PGSs, (HR: 1.03; 95%CI 1.01-1.05, p=0.001) had the weakest and the (HR: 1.17; 95%CI 1.15-1.19, p<0.0001) had the strongest association with an incident fracture. Models that include genome-wide PGSs had higher c-indices than models with restricted PGSs (0.651 versus 0.644) **(Figure 4B)**.

We additionally estimated the OR and HR and corresponding 95% CI for individuals in the bottom 1%, 5%, 10%, and 20% of the PGSs, as compared with the remaining individuals. Results from Cox proportional hazard regression showed that individuals in the bottom 1% distribution of *PGS_FNBMD*_63_, *PGS_FNBMD*_*ldpred*_, and *PGS_TBBMD*_*ldpred*_ had 1.33-, 1.25-, and 1.47-fold increased fracture risk respectively, compared to their corresponding remaining individuals. In contrast, individuals with extreme *PGS_TBBMD*_81_ values did not show a significantly higher risk of fracture. Similar results were observed when applying multiple logistic regression models (**Supplementary Table 2**).

The Cox proportional hazard model showed that, after adjusting for FRAX risk factors available in the UKB testing set, all four PGSs were significantly associated with incident fractures. Out of four PGSs, *PGS_TBBMD*_*ldpred*_ had the strongest association with incident fracture. The HRs of *PGS_FNBMD*_63_, *PGS_TBBMD*_81_, *PGS_FNBMD*_*ldpred*_, and *PGS_TBBMD*_*ldpred*_ for incident fracture were 1.13 (95% CI, 1.11 – 1.15), 1.03 (95% CI, 1.01 – 1.05), 1.11 (95% CI, 1.09 – 1.14), and 1.16 (95% CI, 1.15 – 1.19), respectively. Compared to the FRAX base model, the association between clinical risk factors and incident fracture risk did not attenuate in all four PGS models **(Table 1)**.

**Table 1.** Hazard Ratio of Significant Predictive Variables for Incident Fractures in Models with and without PGSs.

| Variable | Model 1:<br>FRAX Base Model | Model 2:<br>FRAX +<br>*PGS_FNBMD*<sub>63</sub> | Model 3:<br>FRAX +<br>*PGS_TBBMD*<sub>81</sub> | Model 4:<br>FRAX +<br>*PGS_FNBMD*<sub>ldpred</sub> | Model 4:<br>FRAX +<br>*PGS_TBBMD*<sub>ldpred</sub> |
| --- | --- | --- | --- | --- | --- |
|  | HR per 1 unit<br>(95% CI) | HR per 1 unit<br>(95% CI) | HR per 1 unit<br>(95% CI) | HR per 1 unit<br>(95% CI) | HR per 1 unit<br>(95% CI) |
| Age | 1.03 (1.02-1.03) | 1.03 (1.02 – 1.03) | 1.03 (1.02 – 1.03) | 1.03 (1.02 – 1.03) | 1.03 (1.02 – 1.03) |
| Sex (women vs. men) | 2.83 (2.70 - 2.94) | 2.83 (2.70 - 2.94) | 2.83 (2.70 – 2.94) | 2.83 (2.70 – 2.94) | 2.81 (2.63 – 2.94) |
| Body weight | 1.01 (1.01 – 1.01) | 1.01 (1.01 – 1.01) | 1.01 (1.01 – 1.02) | 1.01 (1.01 – 1.02) | 1.01 (1.01 – 1.02) |
| Height | 0.98 (0.98 – 0.99) | 0.98 (0.98 – 0.99) | 0.98 (0.98 – 0.99) | 0.98 (0.98 – 0.99) | 0.98 (0.98 – 0.99) |
| Oral glucocorticoid | 1.10 (0.87 – 1.22) | 1.11 (0.88 – 1.39) | 1.10 (0.86 – 1.37) | 1.09 (0.87 – 1.39) | 1.09 (0.86 – 1.39) |
| Type 1 diabetes | 1.49 (1.30 – 1.69) | 1.48 (1.30 – 1.69) | 1.48 (1.30 – 1.69) | 1.47 (1.28 – 1.69) | 1.46 (1.27 – 1.67) |
| Early menopause | 1.02 (0.97 – 1.08) | 1.02 (0.97 – 1.07) | 1.02 (0.93 – 1.03) | 1.02 (0.93 – 1.03) | 1.02 (0.93 – 1.03) |
| Rheumatoid arthritis | 1.10 (1.01 – 1.19) | 1.10 (1.01 – 1.19) | 1.10 (1.01 – 1.19) | 1.10 (1.02 – 1.20) | 1.10 (1.01 – 1.20) |
| Current smoking | 1.51 (1.43 – 1.59) | 1.50 (1.43 – 1.59) | 1.50 (1.43 – 1.59) | 1.51 (1.43 – 1.59) | 1.51 (1.43 – 1.59) |
| PGS | NA | 1.13 (1.11 – 1.15) | 1.03 (1.01 – 1.05) | 1.11 (1.09 – 1.14) | 1.16 (1.15 – 1.19) |

### Model Evaluation

The fracture discrimination ability of PGSs over clinical risk factors was assessed using the concordance index (c-indices) **(Supplementary Table 3)**. Compared to the base model, models with PGSs showed moderate improvement in discriminating fracture cases and controls. The *PGS_FNBMD*_63_ and the *PGS_TBBMD*_*ldpred*_ improved the discrimination from 0.678 to 0.683 and from 0.678 to 0.686, respectively. In the reclassification analysis, compared to the FRAX base model, the models with *PGS_FNBMD*_63_, *PGS_TBBMD*_81_, *PGS_FNBMD*_*ldpred*_, and *PGS_TBBMD*_*ldpred*_ improved the reclassification of fracture by 2% (95% CI, 1.5% to 2.4%), 0.2% (95% CI, 0.1% to 0.3%), 1.4% (95% CI, 1.3% to 1.5%), and 2.2% (95% CI, 2.1% to 2.4%), respectively. The *PGS_TBBMD*_*ldpred*_ showed the greatest improvement in terms of reclassification. For the model that included *PGS_TBBMD*_*ldpred*_, 395 individuals were correctly reclassified up to the high-risk group, and 325 individuals who did not experience a fracture were correctly reclassified from the high-risk group to the low-risk group. The continuous NRI showed that improvement in fracture reclassification contributed by *PGS_FNBMD*_63_, *PGS_TBBMD*_81_, *PGS_FNBMD*_*ldpred*_, and *PGS_TBBMD*_*ldpred*_ were 11.8%, 2.1%, 7.1%, and 13.2%, respectively (**Table 2**).

**Table 2:**
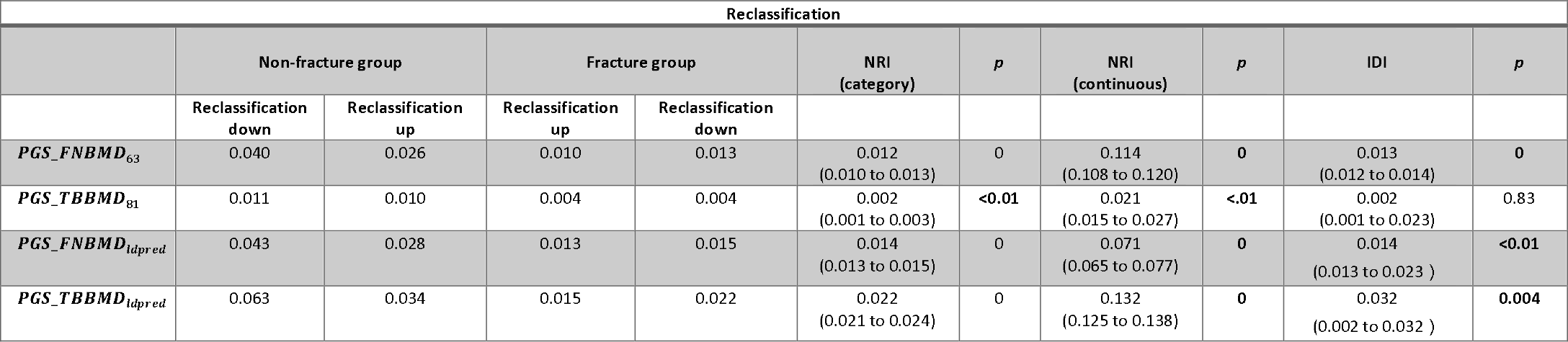
Reclassification Table of 10-Year Osteoporotic Fracture Stratified by Event Status. Results of Reclassification Analysis: Percent of Reclassification Compared with FRAX Base Model.

## Discussion

Early identification of high-risk individuals is crucial in enhancing fragility fracture screening and facilitating preventive interventions [36]. PGS has the advantage that it can be assessed well before any clinical risk factors emerge. As fragility fracture has a sizable heritable component because of its polygenicity nature, utilizing thousands of genetic variants discovered from GWAS to predict risk holds promise for risk stratification and therefore helps facilitate primary prevention.

Prior studies focused mainly on the predictive ability of PGS derived using genome-wide significant SNPs, resulting in mixed findings. This study systematically derived and validated a genome-wide PGS of femoral neck BMD and total body BMD, incorporating information from the entire genome system. To compare the predictive ability of genome-wide PGSs to restricted PGSs, we additionally calculated two previously published PGSs based on 63 femoral neck BMD- and 81 total body BMD-related SNPs, respectively. We quantified the strengths of associations of four PGSs with fracture outcome in 450,000 UKB participants and demonstrated that PGS accurately predicted striking differences in fracture risk. For the total body BMD, our results showed that the LDpred2 approach, which builds a risk prediction model based on the entire genome, yielded better predictive performance than the approach that includes only 81 variants that reached a genome-wide significant level. However, femoral neck BMD-related PGS calculated using the LDpred2 method showed no improvement over the restricted PGS.

Whether including more SNPs would improve the predictive ability of PGS remains controversial. For many phenotypes, genome-wide PGSs outperform those PGSs calculated by using genome-wide significant variants only, in line with the evidence that much of the genetic predisposition of a disease/trait explained by the low-level effect SNPs [37, 38]. However, in some cases, including millions of SNPs with negligible effect size in the polygenic score does not affect the predictions [39-42]. In a prior study, Khera et al. constructed 30 genome-wide PRSs for five common diseases using up to 7 million SNPs. Results show that genome-wide PRSs had lower c-statistics than PRSs based on genome-wide significant SNPs only [43].

In our study, individuals in the top 1% of total body BMD PGS had a HR of 1.47, compared to the remaining individuals. This level of effects may be sufficient to justify the use of PGSs for clinical screening of individuals in order to detect those in the extreme tail, which may be useful for monitoring and preventive treatment. Several studies have investigated the potential for risk scores based on GWAS-level significant variants in improving fracture risk prediction accuracy and reported weak to no evidence for added value from these scores [44-46]. More recently, Lu et al. derived a genome-wide PGS (gSOS) of heel ultrasound measurements (speed of sound) using a statistical learning approach (LASSO) and demonstrated that gSOS was more predictive of major osteoporotic fracture and hip fracture than most clinical risk factors. Additionally, they also derived a FRAX-gSOS and demonstrated that it could refine the risk prediction by employing a positive net reclassification index ranging from 0.024 to 0.072.

However, as a well-used metric of fracture risk that is incorporated into the FRAX algorithm, the genome-wide PGS for BMD has never been studied. In current study, we generated more accurate genome-wide PGSs that can possibly capture a larger proportion of total variance in BMD. BMD-related genome-wide PGSs remained significantly associated with incident fracture risk, even after accounting for FRAX clinical risk factors. Moreover, adding genome-wide PGS to the FRAX clinical risk score has successfully demonstrated significant improvement in predictive accuracy for fracture. The PGS refined risk discrimination and reclassified up to 2% of individuals to a higher or lower fracture risk category. Notably, for total body BMD, in comparison to the restricted PGS, the genome-wide PGS showed significantly better ability in reclassifying individuals who will and will not sustain a fracture.

There are several limitations in the current study worth noting. First, only European ancestry individuals were considered in this study; therefore, the specific PGS calculated here may not have optimal predictive power in other ethnic groups due to different allele frequencies, LD patterns, and effect sizes of common variants across populations of different ethnic backgrounds. Thus, our findings may not generalize to other ethnic groups. Second, due to the limited data availability, we failed to include all 14 clinical risk factors included in FRAX; consequently, a comprehensive evaluation of PGS with complete adjustment was not conducted. Third, the UKB participants were generally younger and healthier than the general population, with a lower incident rate of fracture; this non-random ascertainment is likely to deflate disease prevalence.

In summary, we constructed two genome-wide PGS for BMD based on the UKB dataset and demonstrated that an efficient PGS estimate enables the identification of strata with up to 1.5-fold difference in fracture incidence. This finding definitely calls for personalized screening and prevention strategies that incorporate the PGS information into clinical diagnosis, thus considerably increasing the benefits of population-wide screening programs.

## Data Availability

Data sharing does not apply to this article as no datasets were generated during the current study.

## Acknowledgments

The study was supported by the National Institute of General Medical Sciences under Award Number P20GM121325, and by the National Institute on Minority Health and Health Disparities of the National Institutes of Health under Award Number 1R21MD013681. In addition, the National Supercomputing Institute at the University of Nevada Las Vegas provided facilities for bioinformatical analysis in this study.

