## Supplementary materials for "The Clinical Utility of the BMD-related comprehensive Genome-wide polygenic score in identifying individuals with a high risk of osteoporotic fractures"

**Supplementary Table 1. Participant Characteristics of the UK Biobank Testing Set (N=455,663)**

|  | UKB (N=455,663) | Male  (n=208,271)  (45.71%) | Female  (n=247,392)  (54.29%) |
| --- | --- | --- | --- |
| Age at assessment, yrs. | 56.76 ± 8.02 | 56.99 ± 8.11 | 56.58 ± 7.95 |
| Height (cm) | 168.66 ± 9.25 | 175.84 ± 6.78 | 162.63 ± 6.25 |
| Weight (kg) | 78.16 ± 15.91 | 86.18 ± 14.30 | 71.40 ± 13.95 |
| Current smoker | 47,486 (10.42%) | 25,392 (12.19%) | 22,094 (8.93%) |
| Body mass index (BMI) $(kg/m^{2})$ | 27.39 ± 4.77 | 27.85 ± 4.24 | 27.01 ± 5.14 |
| Fractures in the past 5 years | 47,576 (10.44%) | 19,262 (9.25%) | 28,314 (11.44%) |
| Oral glucocorticoid user | 2,426 (0.53%) | 1,069 (0.51%) | 1,354 (0.55%) |
| Rheumatoid arthritis | 10,964 (2.41%) | 3,612 (1.73%) | 7,352 (2.97%) |
| Type 1 diabetes | 4,336 (0.95%) | 2,517 (1.21%) | 1,819 (0.74%) |
| Menopause before age 45 years | 35,657 (7.83%) | NA | 35,657 (7.83%) |

**Supplementary Table 2. Odds Ratios (OR), Hazard Ratios (HR), and Their Corresponding 95% Confidence Intervals (CI) for Incident Fracture Per 1 SD Decrease in PGS. Results from Multiple Linear Regression and Cox Proportional Hazard Models in the UKB Testing Set.**

| **High PGS definition** | **Reference group** | **Odds ratio (95% CI)** | **P-value** | **Hazard ratio (95% CI)** | **P-value** |
| --- | --- | --- | --- | --- | --- |
| $\boldsymbol{PGS\_FNBMD}_{\boldsymbol{63}}$ | | | | | |
| Bottom 20% of distribution | Remaining 80% | **1.35 (1.28 – 1.41)** | **<0.0001** | **1.21 (1.16 – 1.27)** | **<0.0001** |
| Bottom 10% of distribution | Remaining 90% | **1.39 (1.32 – 1.47)** | **<0.0001** | **1.27 (1.20 – 1.33)** | **<0.0001** |
| Bottom 5% of distribution | Remaining 95% | **1.41 (1.32 – 1.52)** | **<0.0001** | **1.30 (1.22 – 1.39)** | **<0.0001** |
| Bottom 1% of distribution | Remaining 99% | **1.59 (1.39 – 1.85)** | **<0.0001** | **1.33 (1.16 – 1.54)** | **<0.0001** |
| $\boldsymbol{PGS\_FNBMD}_{\boldsymbol{ldpred}}$ | | | | | |
| Bottom 20% of distribution | Remaining 80% | **1.32 (1.25 – 1.37)** | **<0.0001** | **1.20 (1.15 – 1.25)** | **<0.0001** |
| Bottom 10% of distribution | Remaining 90% | **1.35 (1.28 – 1.43)** | **<0.0001** | **1.20 (1.14 – 1.27)** | **<0.0001** |
| Bottom 5% of distribution | Remaining 95% | **1.41 (1.32 – 1.54)** | **<0.0001** | **1.25 (1.16 – 1.35)** | **0.002** |
| Bottom 1% of distribution | Remaining 99% | **1.59 (1.35 – 1.85)** | **<0.0001** | **1.25 (1.06 – 1.47)** | **<0.0001** |
| $\boldsymbol{PGS\_TBBMD}_{\boldsymbol{81}}$ | | | | | |
| Bottom 20% of distribution | Remaining 80% | 1.02 (0.98 – 1.08) | 0.27 | 1.02 (0.98 – 1.07) | 0.26 |
| Bottom 10% of distribution | Remaining 90% | 1.02 (0.96 – 1.09) | 0.46 | **1.06 (1.01 – 1.12)** | **0.05** |
| Bottom 5% of distribution | Remaining 95% | 1.08 (0.99 – 1.16) | 0.09 | **1.13 (1.04 – 1.22)** | **0.002** |
| Bottom 1% of distribution | Remaining 99% | 1.08 (0.90 – 1.27) | 0.41 | 1.14 (0.96 – 1.35) | 0.14 |
| $\boldsymbol{PGS\_TBBMD}_{\boldsymbol{ldpred}}$ | | | | | |
| Bottom 20% of distribution | Remaining 80% | **1.47 (1.43 – 1.54)** | **<0.0001** | **1.28 (1.23 – 1.33)** | **<0.0001** |
| Bottom 10% of distribution | Remaining 90% | **1.59 (1.51 – 1.69)** | **<0.0001** | **1.31 (1.25 – 1.37)** | **<0.0001** |
| Bottom 5% of distribution | Remaining 95% | **1.69 (1.59 – 1.82)** | **<0.0001** | **1.36 (1.28 – 1.45)** | **<0.0001** |
| Bottom 1% of distribution | Remaining 99% | **1.89 (1.64 – 2.17)** | **<0.0001** | **1.47 (1.30 – 1.67)** | **<0.0001** |

**Supplementary Table 3. Concordance Index and the Corresponding 95% Confidence Intervals of Predicted and Observed Fracture Risk for the Model with and without PGS.**

|  | C-index | 95% CI | P-value |
| --- | --- | --- | --- |
| Model 1  (Base model) | 0.663 | 0.658 – 0.668 | NA |
| Model 2  (Base model + $\boldsymbol{PGS\_FNBMD}_{\boldsymbol{63}}$) | 0.668 | 0.662 – 0.672 | <0.001 |
| Model 3  (Base model + $\boldsymbol{PGS\_TBBMD}_{\boldsymbol{81}}$) | 0.663 | 0.660 – 0.672 | <0.001 |
| Model 4  (Base model + $\boldsymbol{PGS\_FNBMD}_{\boldsymbol{ldpred}}$) | 0.665 | 0.661 – 0.672 | 0.007 |
| Model 5  (Base model + $\boldsymbol{PGS\_TBBMD}_{\boldsymbol{ldpred}}$) | 0.668 | 0.661 – 0.673 | <0.001 |

| **A.** | **B.** | **C.** |
| --- | --- | --- |
| 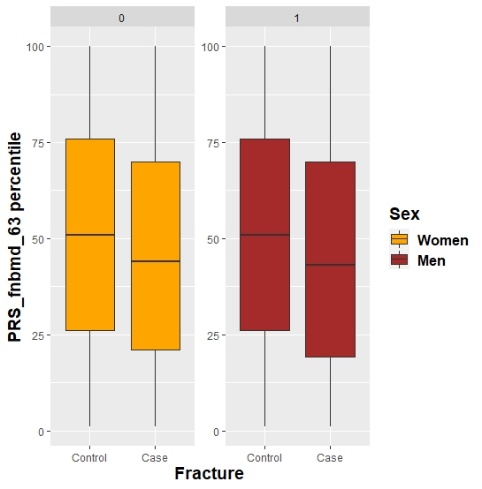 | 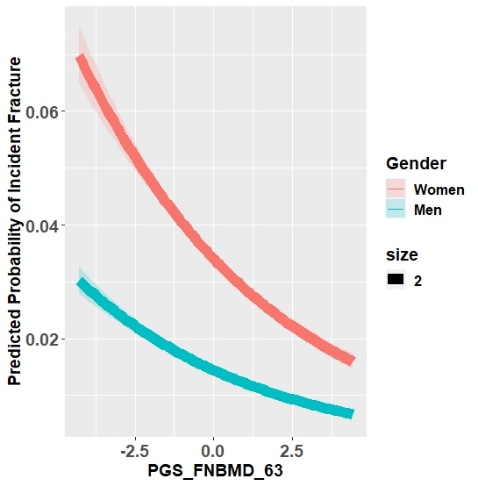 | 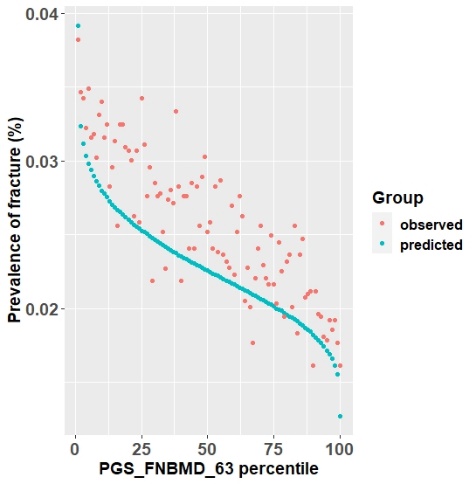 |
| 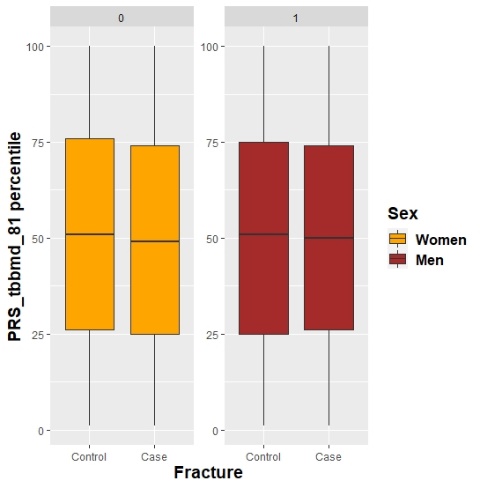 | 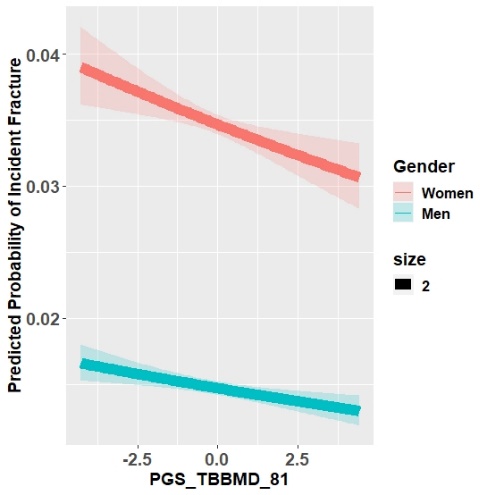 | 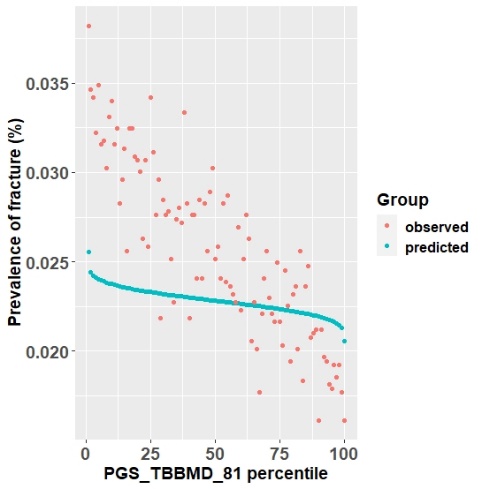 |
| **(A)** PGSs percentile among fracture cases versus controls in the UK Biobank testing set. Within each boxplot, the horizontal lines reflect the median, the top and bottom of each box reflect the interquartile range, and the whiskers reflect the maximum and minimum values within each group. **(B)** Predicted Probability of Incident Fracture by PGSs: Risk gradient for fractures according to the PGS percentiles. 100 groups of the testing dataset were derived according to the percentile of each of the four PGSs. **(C)** Predicted versus Observed prevalence of incident fracture according to PGS percentiles. | | |

**Supplementary Figure 1. Risk for Incident Fracture According to Restricted PGSs.**
